# 9.4 Tesla MRI in focal epilepsy patients with high-resolution surface-based profiling of focal cortical dysplasias

**DOI:** 10.64898/2026.04.02.26349812

**Authors:** Cornelius Kronlage, Pascal Martin, Benjamin Bender, Gisela E. Hagberg, Jonas Bause, Joana RA Loureiro, Mathilde Ripart, Sophie Adler, Konrad Wagstyl, Holger Lerche, Niels KN Focke, Klaus Scheffler, Esther Kuehn

## Abstract

1.

**Background:** The detection of subtle epileptogenic lesions such as focal cortical dysplasias (FCDs) is a clinical challenge in the management of drug-resistant focal epilepsy (DRFE). Ultra-high field (UHF) MRI offers increased signal-to-noise ratios and spatial resolution compared to 3 Tesla (T) MRI and may improve diagnostic yield. Here, we present a 9.4T MRI cohort study of patients with DRFE.

**Methods:** We recruited n=21 DRFE patients (with 3T-MRI findings: 2 positive, 3 equivocal, 16 negative) undergoing presurgical workup, and n=20 healthy controls for 9.4T MRI (0.8 mm isotropic MP2RAGE, slabs of 0.375 × 0.375 × 0.8 mm T2*-weighted GRE) and 3T MRI (MP2RAGE, FLAIR) acquisitions. Visual review for possible epileptogenic lesions was performed by clinical experts. For histopathologically confirmed FCD lesions, we extracted surface-based quantitative features (cortical thickness, qT1, FLAIR, T2*, and QSM values) across cortical depths and distances from the lesion centre and performed high-resolution cortical profiling of 9.4T T2* values.

**Results:** No new epileptogenic lesions were visually identified at 9.4T in 3T MRI negative patients. In the two patients with histopathologically confirmed lesions, the FCD IIb lesions were visible with distinct qualitative and quantitative features at both field strengths. One of these FCD IIb showed a focal cortical T2* reduction at 9.4T that could here be quantified via automated cortical profiling, consistent with the previously described “black line sign”.

**Conclusion:** 9.4T MRI findings in epileptogenic lesions underlying DRFE are consistent with those on 3T MRI. While additional lesions were not identified in patients with negative 3T MRI, higher resolution T2*-weighted sequences can reveal a feature not seen at 3T: Cortical profiling of FCDs highlights the black line sign and can possibly help refine surgical or ablation targeting for some FCDs. Further optimization of UHF protocols and analysis methods on larger cohorts may reveal clinically applicable diagnostic benefits.

**Key Points:**

- 9.4T MRI shows focal cortical dysplasia (FCD) IIb with distinct qualitative and quantitative features that are consistent with 3T MRI.
- High-resolution quantitative T2* maps at 9.4T may provide additional information for defining resection or ablation targets in FCDs.
- Without using parallel transmit technology, artefacts in the temporal lobes pose a limitation of 9.4T MRI in presurgical epilepsy workup.

## 2. Introduction

In drug-resistant focal epilepsy (DRFE), epilepsy surgery can achieve seizure freedom by resection or ablation of the epileptogenic zone. Pinpointing epileptogenic lesions on MRI is therefore critical. However, lesions are often inconspicuous, may escape detection by MRI and are only identified on histopathology.^1,2^ Resection of MRI-diagnosed lesions is associated with better clinical outcomes,^3^ whereas MRI-negative patients may need to undergo additional invasive investigations like intracranial electroencephalography (EEG). Furthermore, as minimally invasive procedures such as laser interstitial thermal therapy (LITT) and radiofrequency ablation (RFA) are becoming more common^4^, and limited resections of bottom-of-sulcus FCDs have been reported to be effective^5,6^, high-resolution characterization of epileptogenic lesions is increasingly necessary. Improving the sensitivity, specificity and spatial precision of MRI to detect epileptogenic lesions can therefore be expected to translate into direct clinical benefit in the management of DRFE patients.

Focal cortical dysplasias (FCDs) represent one of the most common pathologies in DRFE. The features of FCDs vary according to their subtype. FCD Type II lesions, the most common subtype, are characterized on MR imaging by cortical thickening, subcortical T2/FLAIR hyperintensity extending to the ventricles (transmantle sign) and blurring of the interface of grey-matter (GM) and white matter (WM).^7^ They can be small, subtle and hard to detect visually even for expert readers (with a detection rate of, e.g., 68%).^8^ Quantification of these radiological features by computational methods is possible^9,10^ and relevant as they serve as inputs for (deep learning) detection algorithms that may improve diagnostic yield in previously MRI-negative cases.^11,12^

Ultra-high field (UHF) MRI is defined as MR-based imaging at magnetic field strengths of 7 Tesla (T) or more. Compared to 1.5 or 3T MRI, UHF-MRI enables measurements with a higher signal-to-noise ratio (SNR),^13^ which can be traded for higher spatial resolution and/or shorter measurement duration. Furthermore, UHF-MRI facilitates the acquisition of some contrasts like T2*-weighted images. On the other hand, certain physical properties can translate into disadvantages: Higher field strengths and the associated higher Larmor frequencies relate to reduced radiofrequency (RF) uniformity – resulting in artefacts – and increased RF power deposition – requiring adaptations of protocols.^14^

Consequently, UHF-MRI can be expected to improve diagnostic sensitivity in DRFE. Small structural features may be captured with higher fidelity, e.g., blurring of the GM/WM border in FCDs or loss of hippocampal structure in hippocampal sclerosis. A few studies have employed 7 T MRI for lesion detection in DRFE, reporting diagnostic gains (i.e., the ratio of patients with new relevant findings to previously MRI-negative patients studied) of around 20-30%.^15–21^ As a novel feature detectable by UHF-MRI, the term “black line sign” has been coined to describe a focal cortical T2* hypointensity in FCD II.^22,23^ Complete resection of the black line sign has been shown to be associated with better surgical outcome. While 7 T MRI is becoming more widely available, only a few human 9.4T systems have been installed worldwide so far. Given SNR increases with field strength supra-linearly,^13^ 9.4T MRI may offer yet another substantial gain in resolution and diagnostic power. In-vivo human measurements at even higher field strengths of, e.g., 11.7 T^24^ are possible and may become more relevant in the future.

To the best of our knowledge, apart from preliminary data from this project,^25^ to date, only one case report of a focal epilepsy patient scanned at 9.4T has been published.^26^ In addition, a protocol that includes 9.4T MRI on 7 T MRI-negative patients has been published and the study is ongoing.^27^

Here, we present a cohort of DRFE patients and healthy controls examined using in-vivo 9.4T MRI with standardized 3T MRI acquired for comparison. We investigated whether 9.4T MRI (1) increases diagnostic yield, i.e. reveals epileptogenic lesions in 3T MRI negative patients; (2) reveals additional features or more intra-lesional detail particularly in FCDs; (3) enhances effect sizes of quantitative features characteristic of FCDs. These findings provide insights into the potential of UHF-MRI to improve cortical profiling of FCDs and lesion detection in DRFE.

## 3. Methods

### 3.1. Inclusion and exclusion criteria

N=21 adult (> 18 years old) patients with drug-resistant focal epilepsy undergoing presurgical diagnostic workup and n=20 healthy controls were recruited to participate in this observational diagnostic study. Exclusion criteria were the inability to provide informed consent, contraindications for UHF MRI (such as implants or claustrophobia but also tattoos), and pregnancy or breastfeeding.

Recruitment and MRI examinations were conducted between March 2015 and October 2018. The study was approved by the local ethics committee (Tübingen University, reference number 390/2014BO1), participants and patients provided informed consent. In line with the ethics approval for this study, 9.4T MRI data was not used to inform clinical treatment decisions. The subject IDs used in this manuscript (e.g., P012) are pseudonyms assigned exclusively for the purposes of this study, no information linking these identifiers to patients’ identities or medical records was accessible to anyone outside the research group.

### 3.2. Demographic and clinical data

Demographic and clinical data were collected in the epilepsy surgery multidisciplinary team meetings at the University Hospital Tübingen. Postoperative outcomes and histopathology for surgically treated patients were gathered from the hospital electronic health record.

Patients had a median age of 27 years (range 18-51), 11 were female, 10 male. Controls had a median age of 28 years (range 21-50 years, 7 female, 13 male. For detailed information, see table 1. Most notably, 2/21 patients (10%) had a clear finding on 3T MRI (2 FCD – P017 and P026). Another 3/21 (14%) had a suspected epileptogenic lesion (2 FCD – P014, P016; 1 hippocampal sclerosis – P009) reported in at least one but not each of the clinical MRI examinations. The remaining patients (76%) were defined as 3T MRI negative. With respect to non-invasive video-EEG, which was conducted in all patients, seizures were recorded in all but 1 patient. 13/21 patients (62%) underwent 18F-desoxyglucose positron emission tomography (FDG-PET), and 2/13 had localizing concordant findings. Intracranial EEG was performed in 6/21(29%). 3/21 patients (14% - P014, P017, P026) underwent surgical resections, histopathology revealed FCD IIb in 2/3 (P017, P026) and only fragmented tissue without evidence of pathology in 1/3 (P014). The operated patients with FCD2b (P017, P026) had good clinical outcomes (Engel class I).

**Table 1:** Patient demographic and clinical information, L: left, R: right, F: frontal, I: insular, T: temporal, P: parietal, O: occipital, HS: hippocampal sclerosis, FCD: focal cortical dysplasia, surgical outcome specified as Engel class (I-IV), *years specified in age groups (min. 5 years range) for de-identification

| id | age<br>[years*<br>] | sex | age at<br>onset of<br>epilepsy<br>[years*] | 3T MRI<br>diagnosis | suspected<br>pathology<br>on 3T MRI | clinical<br>hypothesis<br>(lobar) | FDG PET<br>finding/<br>localization | neuro-psychology | non-invasive<br>video-EEG | invasive<br>EEG<br>localization | surgery<br>performed | contraindication<br>to surgery | histopathology | postsurgical<br>outcome (Engel<br>class) |
| --- | --- | --- | --- | --- | --- | --- | --- | --- | --- | --- | --- | --- | --- | --- |
| P003 | 18-25 | f | 11-15 | none | n/a | R F | n/a | R F-T | R F | n/a | 0 | n/a | n/a | n/a |
| P004 | 36-40 | f | 0-5 | none | n/a | R F-I | normal | R F-T | R F-I | R F/I | 0 | multifocal | n/a | n/a |
| P005 | 31-35 | m | 16-20 | none | n/a | L T | n/a | L F-T | L T | n/a | 0 | n/a | n/a | n/a |
| P006 | 26-30 | f | 16-20 | none | n/a | R T | normal | non localizing | R T | n/a | 0 | n/a | n/a | n/a |
| P008 | 46-50 | f | 31-35 | none | n/a | R T | normal | L F-T | R T | n/a | 0 | eloquent area | n/a | n/a |
| P009 | 51-55 | f | 26-30 | suspicion | HS R | L/R T | n/a | L/R F-T | L/R T | n/a | 0 | n/a | n/a | n/a |
| P010 | 41-45 | m | 11-15 | none | n/a | L T | n/a | L F-T | L/R F-T | n/a | 0 | n/a | n/a | n/a |
| P011 | 18-25 | m | 16-20 | none | n/a | L/R F | normal | L/R F | L/R F | n/a | 0 | n/a | n/a | n/a |
| P012 | 18-25 | m | 11-15 | none | n/a | L T-P | normal | L F | L F-T-P | L T-P | 0 | eloquent area | n/a | n/a |
| P013 | 26-30 | f | 16-20 | none | n/a | L/R T | L T | L F-T | L/R T | L+R T | 0 | multifocal | n/a | n/a |
| P014 | 31-35 | f | 0-5 | suspicion | FCD | L C | L F | L F-T | L F | L F | 1 | n/a | fragmented<br>tissue, no FCD | not classifiable |
| P015 | 18-25 | f | 11-15 | none | n/a | L/R F | normal | L/R F | L/R F | n/a | 0 | n/a | n/a | n/a |
| P016 | 26-30 | f | 6-10 | suspicion | FCD | R I, L O | n/a | F | no seizures<br>recorded | n/a | 0 | n/a | n/a | n/a |
| P017 | 18-25 | f | 0-5 | positive | FCD | R F | n/a | F | R F | n/a | 1 | n/a | FCD IIb | I (6 years) |
| P018 | 26-30 | m | 11-15 | none | n/a | L F | normal | L/R F | L F | n/a | 0 | n/a | n/a | n/a |
| P021 | 18-25 | f | 11-15 | none | n/a | L F | n/a | L/R F | L F | n/a | 0 | n/a | n/a | n/a |
| P022 | 18-25 | m | 6-10 | none | n/a | R T-P-O | normal | R T-P | R T-P-O | R T-P-O | 0 | eloquent area | n/a | n/a |
| P023 | 18-25 | m | 0-5 | none | n/a | R F-C-T | normal | R F-T | R F-T | n/a | 0 | n/a | n/a | n/a |
| P024 | 31-35 | m | 16-20 | none | n/a | R T | normal | R F-T | R F-T | n/a | 0 | n/a | n/a | n/a |
| P025 | 26-30 | m | 11-15 | none | n/a | L/R T | normal | R F-T | L/R T | n/a | 0 | n/a | n/a | n/a |
| P026 | 46-50 | m | 31-35 | positive | FCD | L F | n/a | L F-T | L F | L F | 1 | n/a | FCD IIb | I (min. 2 years) |

In some cases with equivocal or negative 3T MRI, ancillary data may have allowed to ascertain and localize lesions but was overall inconclusive, as outlined below:

Patient P014 had an electroclinical left frontal/central hypothesis and was initially MRI-negative. FDG-PET demonstrated focal hypometabolism in the left precentral gyrus, where suspicion of an FCD was raised in a repeat 3T MRI. Invasive EEG confirmed this seizure onset. Resection was performed. Histopathology showed fragmented tissue without evidence of an FCD. The patient discontinued medication postoperatively and continued to have seizures, rendering the outcome uninterpretable.

Patient P016 showed right insular and left occipital T2/FLAIR abnormalities – considered possible FCDs – on 3T MRI. No seizures were recorded during video-EEG monitoring, and psychogenic non-epileptic seizures were later discussed.

Patient P009 had right hippocampal sclerosis on 3T MRI, but seizure onset could not be clearly lateralized on video-EEG. Surgery was not pursued.

Four additional patients with negative 3T MRI underwent invasive EEG. In P004, P012, and P013, numerous seizures were recorded and multiple seizure onsets identified in each case, precluding surgery. In P022, a single seizure with mesial temporo-occipital onset was recorded, but surgery was contraindicated due to proximity to the optic radiation and primary visual cortex.

### 3.3. MRI data acquisition

#### 3.3.1. 9.4T MRI

9.4T MRI scans were acquired with a human whole-body 9.4T MRI scanner (Siemens Healthineers, Erlangen, Germany) located at the Department for High-Field Magnetic Resonance of the Max Planck Institute for Biological Cybernetics, Tuebingen. A custom built head coil with a 16 element dual row transmit array and a 31-element receive array^28^ was used. MRI sequences included: (1) Whole brain 0.8 mm isotropic MP2RAGE (Magnetization Prepared 2 Rapid Acquisition Gradient Echoes^29^) (TE 2.3 ms, TR 6 ms, TI1 900 ms, TI2 3500 ms, FA1 4°, FA2 6°, volume TR: 9 s); (2) a T2*-weighted mono-polar multi-echo GRE with 0.375 × 0.375 × 0.8 mm^3^ voxel size (TE 6–30 ms in steps of 6 ms, TR 35 ms, FA 11°)) acquired in slabs positioned according to the suspected lesions or clinical hypotheses. 9.4T MP2RAGE UNI and T1 maps were reconstructed offline with correction for excitation flip angle and inversion efficiency deviations.^30^ Multi-echo GRE data were processed as described previously^31,32^ to generate T2* and QSM maps.

#### 3.3.2. 3T MRI

3T MRI scans were collected with a MAGNETOM Prisma scanner (Siemens Healthineers, Erlangen, Germany) with a 64 channel head coil. The following data were acquired: Each whole brain 1.0 mm isotropic (1) T1-weighted MPRAGE (Magnetization-Prepared Rapid Gradient-Echo) (TE 2.98 ms, TR 2300 ms, TI 900 ms, FA 9°, (2) MP2RAGE (TE 2.98 ms, volume TR 5000 ms, TI1 700 ms, TI2 2500 ms, FA1 4°, FA2 5°); (3) T2-FLAIR weighted sequence (Sampling Perfection with Application optimized Contrasts using different flip angle Evolution-Fluid-Attenuated Inversion Recovery, T2-SPACE FLAIR) (TE 388 ms, TR 5000 ms, TI 1800 ms, FA 120°).

### 3.4. Visual evaluation

9.4T and 3T MR images were reviewed by a neuroradiologist (B. B.) and two neurologists (C. K., P. M.) with expertise in imaging of focal epilepsies with respect to the presence and appearance of possible epileptogenic lesions or other incidental findings. Reviewers were not blinded and had access to clinical information. Qualitative findings were recorded.

### 3.5. Lesion masks

To guide the extraction of features by surface-based morphometry, lesions were manually labeled. For operated patients, pre-operative (9.4T and 3T) as well as clinical post-operative images were used to define lesion masks using the FreeView software (C. K. and P. M.).

### 3.6. Surface-based analysis

#### 3.6.1. Segmentation and cortical surface reconstruction

MP2RAGE ‘uniform’ T1-weighted images contain salt and pepper noise outside the brain that impedes processing with pipelines adapted to conventional T1-weighted data such as Freesurfer. Therefore, 9.4T / 3T MP2RAGE images were denoised by calculating ‘robust’ uniform T1w images based on the GRE_TI1_ and GRE_TI2_ data^33^ with the parameter beta set to 0.4. As the denoising reintroduces some bias field artifacts, N4 bias field correction^34^ was applied subsequently.

The denoised uniform 9.4T and 3T MP2RAGE images, respectively, were then used for segmentation and surface reconstruction using the Freesurfer recon-all pipeline,^35^ version 7.4.1, set to operate at native sub-millimetric resolution (‘-hires’ flag) and to use synthstrip^36^ for improved skull stripping quality.

#### 3.6.2. Surface-based features and lesion profiles

The 3T MPRAGE T1w and FLAIR volumes were coregistered (using Freesurfer’s ‘mri_coreg’ utility with 12 degrees of freedom), resampled to the uniform 3T MP2RAGE volumes, N4 bias field corrected, and intensity normalised (by linearly rescaling so that the medians of white matter and cortex voxels are 1 and 0, respectively – similar to a method described previously for FLAIR^37^).

The 9.4T GRE_TE1_ volumes were coregistered to the 9.4T MP2RAGE volumes (using mri_coreg as well). This registration was used to align and resample T2*- and QSM-maps accordingly.

All available volume data (at 3T: uniform MP2RAGE, T1-maps, normalised MPRAGE, normalised FLAIR; at 9.4T: uniform MP2RAGE, T1-maps, T2*-maps, QSM-maps) were projected onto the respective cortical surface mesh at six depths ranging from –1.0 to –6.0 mm from the pial surface in 1.0 mm increments. Cortical thickness was re-estimated without the Freesurfer default upper limit of 5.0 mm.

#### 3.6.3. Registration to a template space

Surface meshes were coregistered to a bilaterally symmetric template (xhemi, fsaverage_sym)^38^ to enable per-vertex comparisons between subjects. All surface features were mapped to this template space.

Similarly, lesion masks were sampled onto the respective subject’s cortical surface mesh and then to the Freesurfer template. To avoid a bias towards one field strength, the set of template vertices identified as lesional based on both 3T and 9.4T reconstructions (i.e., the intersection) was used for further analysis. Similarly, the per-vertex geodesic distance to the lesion centroid (lesional vertex with the smallest distance to the center of mass of all lesional vertices) was computed along the pial surface for both 3T and 9.4T surface meshes and averaged on the template. The lesion masks in the left-right symmetric template space also defined homotopic regions on the contralateral hemisphere for patients and in controls.

#### 3.6.4. Analysis and statistics

Surface-based features were compared between 9.4T and 3T for the n=2 histologically verified FCDs. Vertex-wise data was aggregated by computing the median either in the entire lesion mask (for comparisons across cortical depth) or in bins with equal numbers of vertices in increasing geodesic distance from the lesion mask centroid. Features were normalized by z-scoring (subtraction of mean and division by standard deviation) with respect to the distribution of healthy control subjects. Due to the limited sample size of histopathologically confirmed FCDs (n=2), we did not compute statistical comparisons on this data.

#### 3.6.5. High-resolution profiling of T2* data

To characterise the FCDs in detail in the high resolution (0.375 mm in-plane) 9.4T quantitative T2* maps with respect to the “black line sign”, an additional targeted analysis was performed: The Freesurfer cortical segmentation within the manually labelled lesion and in the surrounding cortex up to a geodesic distance of 100 mm from the lesion centroid was processed using laynii^39^ to assign each voxel to a cortical layer (equi-volumetric layering, with an arbitrarily chosen number of 10 bins) and to a cortical column (average column volume of 40 mm^3^). Column-wise profiles of T2* in different cortical depths were extracted. Based on exploratory analyses and visual verification that columns were selected from anatomically appropriate locations, a profile was classified as consistent with a “black line sign” if had a T2* value < 30 ms in layer 5 and T2* > 30 ms in layer 2. The average profile of columns meeting these criteria was extracted and used as a target profile. For each column, the root mean square error (RMSE) between its T2* profile and the target profile was calculated as a measure of similarity, log-transformed for visualization, and overlaid onto the T2* maps as well as plotted against geodesic distance from the lesion centroid in a scatterplot.

### 3.7. Data and code availability

The healthy volunteer 9.4T MP2RAGE data has been published as part of a larger dataset^40^. It is currently not possible to make the patient imaging data openly accessible due to data protection regulations as specified in the ethics application approved by the local ethics committee (Tübingen University, reference number 390/2014BO1). The code used for analysis is available at https://github.com/ckronlage/3T9T_profiling_pub.

## 4. Results

### 4.1. Visual evaluation of MR images

In the n=16 MRI negative patients, no new epileptogenic lesions concordant with clinical hypotheses were identified upon visual inspection by 3 clinical specialists (all co-authors).

Two histologically verified FCD lesions that were visible at 3T (patients P017 and P026) were clearly discernible at 9.4T (see figure 1, 2). Both lesions were characterized by increased cortical thickness, apparent on both 3T and 9.4T T1-weighted images. In one of the lesions, blurring of the GM/WM border was visible, which was more conspicuous in 9.4T MP2RAGE. Subcortical T2/FLAIR hyperintensity (transmantle sign) in the 3T data was clearly visible in patient P017 and less apparent in patient P026. A similar hyperintensity was visible in the 9.4T GRE_TE1_ magnitude image but less visible in the T2* map in patient P017 (see figure 1). Additionally, a subtle intra-cortical region of decreased T2* values was identified.

**Figure 1:**
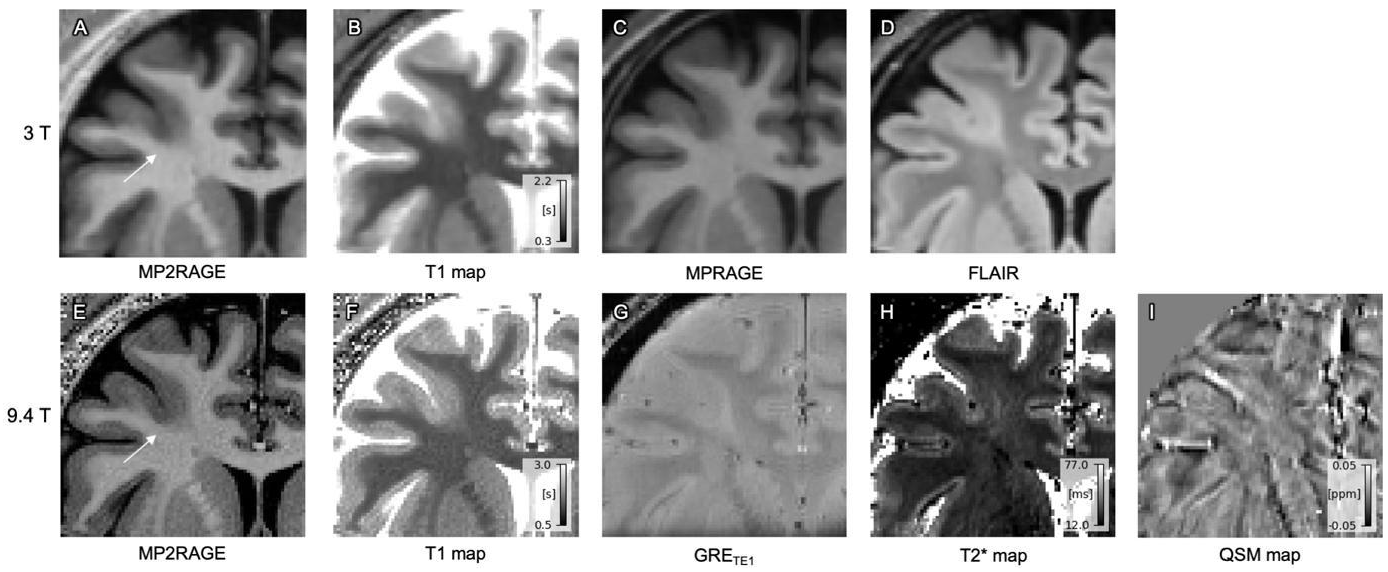
FCD IIb lesion in patient P017. Coronal slices, comparison of 3T data (top row, A-D) and 9.4T data (bottom row, E-I). Thickening of the cortex, blurring of the GM/WM interface and subcortical T2/FLAIR hyperintensity (transmantle sign) are visible on the 3T images as typical imaging features of an FCD. Increased cortical thickness and reduced sharpness of the gray/white matter border are discernible on the 9.4T images as well. The GRE_TE1_ shows subcortical hyperintensity, correspondingly, there are higher T2*-values. A stripe of decreased T2* in the cortex in the center of the lesion (possibly corresponding to the ‘black line sign’ described previously^22,23^) is seen. The QSM shows heterogeneous signal without clear contrast between the lesion and neighbouring areas or GM and WM.

**Figure 2:**
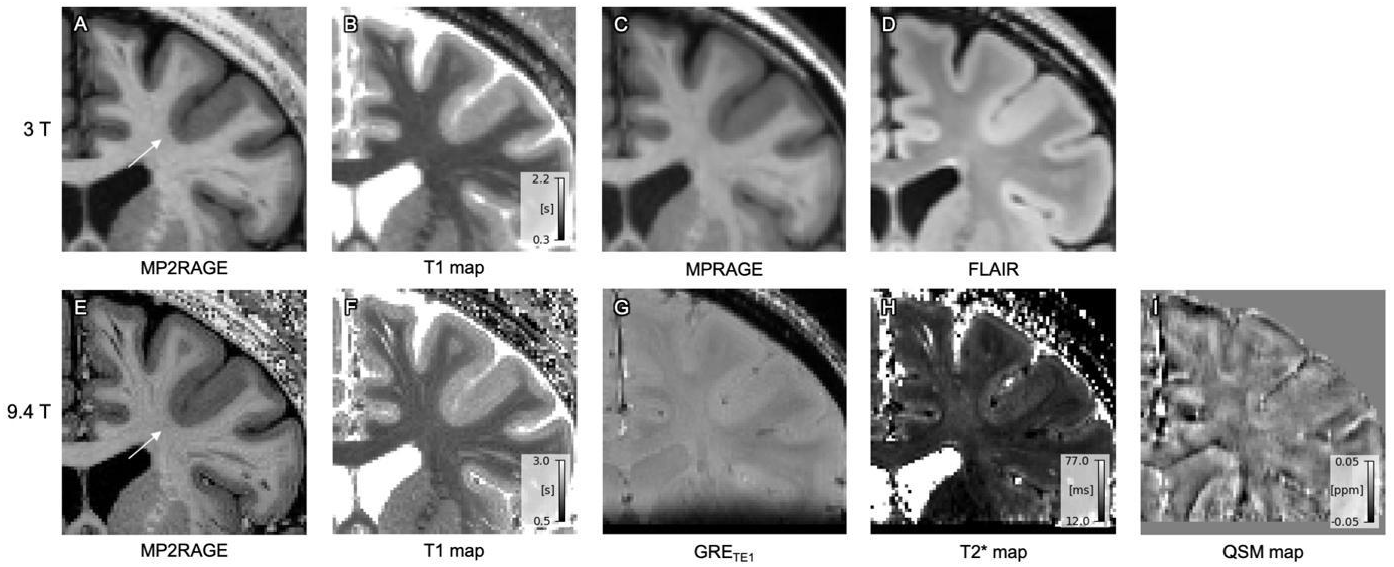
FCD IIb lesion in patient P026. Coronal slices, comparison of 3T data (top row, A-D) and 9.4T data (bottom row, E-I). On the 3T images, thickening of the cortex is visible; in the FLAIR image, a subtle subcortical FLAIR hyperintensity can be suspected. There is no blurring of the GM/WM border. In the 9.4T data, the same imaging features (cortical thickening without blurring) are apparent. The GRE_TE1_ and T2* data do not show a subcortical signal comparable to the 3T FLAIR. There is no clear alteration of QSM in the lesion.

In the patient with suspected right hippocampal sclerosis (see supplementary figure 1), hippocampal volume loss was visible on the 3T as well as on 9.4T T1-weighted images. There was no clear T2/FLAIR hyperintensity visible at 3T3T. The findings were not discernible on 9.4T T2*-weighted GRE images or derived T2* maps.

In patient P016 with suspected FCDs in the posterior right insula and the left occipital gyrus, but without seizures recorded on video-EEG, there were no qualitatively new findings at 9.4T (see supplementary figure 2).

In patient P014 with suspected FCD in the left precentral gyrus reported in one of the preoperative MRI with inconclusive histology and postoperative outcome, imaging findings were neither apparent at 3T nor in the 9.4T MR images.

### 4.2. 9.4T MR image quality

All 9.4T MP2RAGE volumes contained minor artefacts in the lower temporal lobes. Three T1w volumes (from two patients and one control) contained more pronounced artefacts impeding delineation of entire temporal cortical gyri and sulci (see examples in supplementary figure 3).

### 4.3. Surface-based profiling of histologically verified FCDs

Two patients, P017 and P026, had histologically verified FCDs. Surface reconstructions in the lesion areas appeared visually accurate for both field strengths, and showed high agreement (see figure 3A-C, E-G). Estimates of cortical thickness in the lesion ROI were comparable between 3T and 9.4T MR images (numerically slightly larger for 9.4T compared to 3T data for P017, and slightly reduced for P026, median 3.84 vs. 3.94 mm and 3.07 vs. 3.02 mm, respectively, see figure 3D,H). For both 3T and 9.4T MR images, cortical thickness values were clearly above average compared to controls when sampled at homotopic regions (see figure 3D,H).

**Figure 3:**
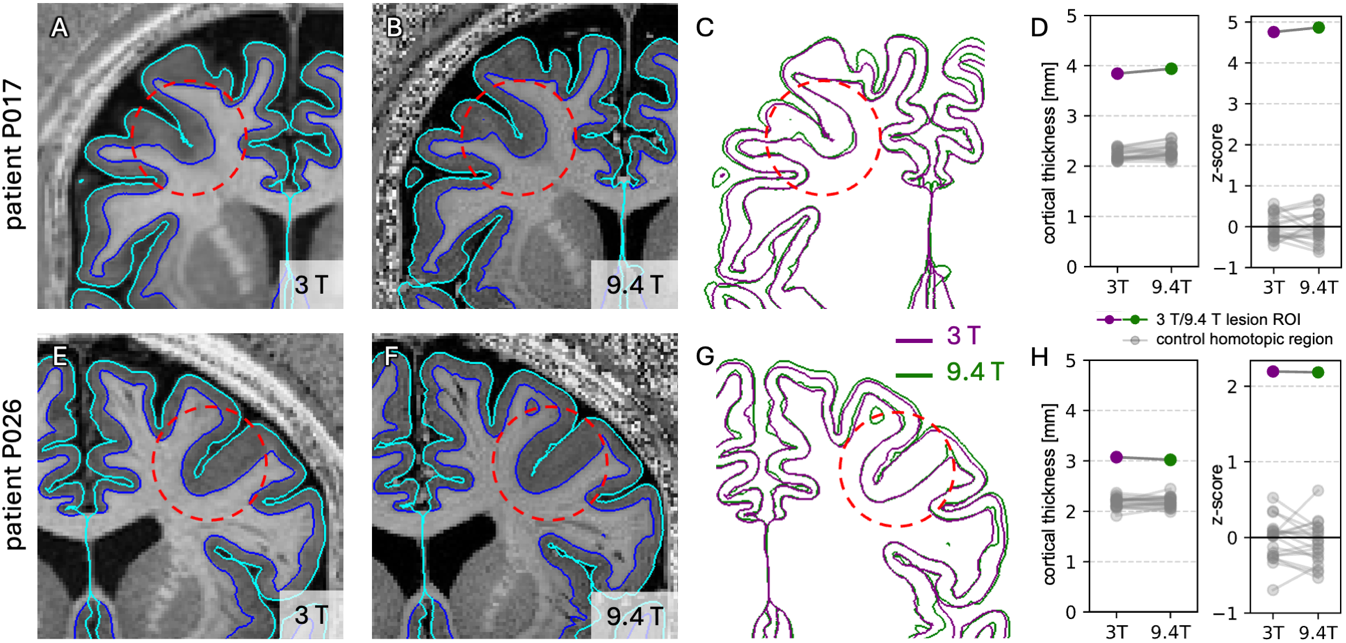
Cortical surface reconstructions overlaid on coronal slices of MP2RAGE uniform volumes (pial surface: cyan, GM/WM boundary: blue), top row (A-D) patient P017, bottom row (E-H) patient P026. The respective lesion areas are highlighted with red circles. (A, E) 3T images, (B, F) 9.4T images, (C, G) overlay of coregistered surface outlines, (D, H) plot of median lesion cortical thickness; raw values left, z-score normalized in reference to healthy controls (plotted in gray) on the right. Visually, there are few differences between reconstructions in the lesion areas.

Both lesions show a distinct depth-dependent profile with regards to some but not all extracted features (see figure 5). Specifically, increased normalized FLAIR intensity, increased quantitative T1 (qT1) values, and increased T2* values at 3-6 mm of sub-pial depth were distinctive features. In contrast, there was no clear effect for estimates based on quantitative susceptibility mapping (QSM) within and below the lesion area. A direct numerical comparison between 3T and 9.4T acquisitions of qT1 values was possible (based on MP2RAGE acquisitions). For qT1 values, no difference in effect sizes between 3T and 9.4T MR images was apparent (see figure 4A). However, statistical tests could not be calculated as only two patients were analyzed.

**Figure 4:**
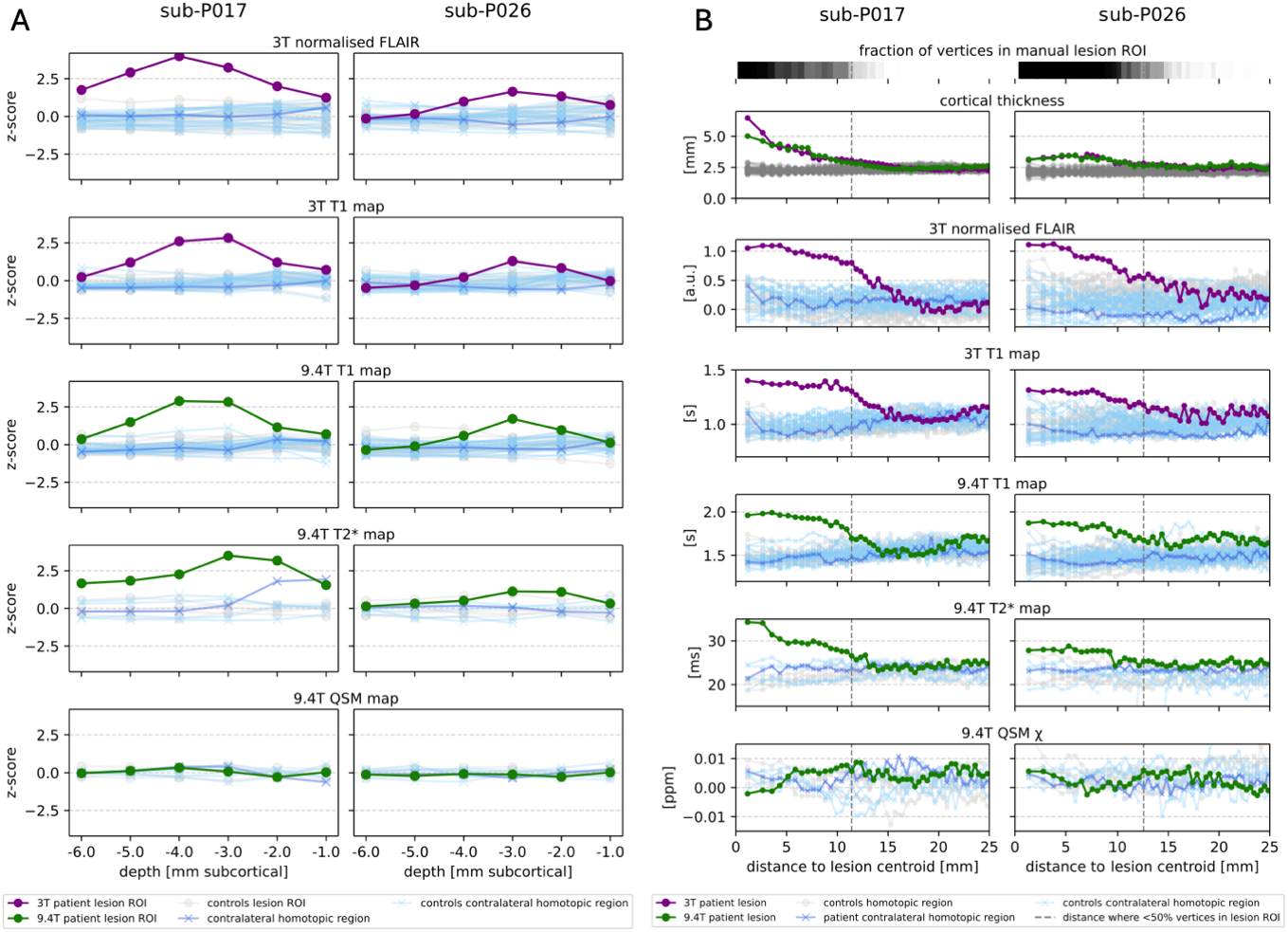
Vertical and horizontal quantitative profiles of two histologically verified FCDs across magnetic field strengths of 3T and 9.4T. For comparison, features in the contralateral homotopic region of patients are shown with crosses in blue, data from controls in light grey and blue respectively. (A) For each feature, the median z-scored value (relative to controls) in the lesion ROI is plotted against sub-pial depth. It is visible that both lesions show FLAIR-hyperintensity, higher T1 values, higher T2* values and no clear changes in susceptibility. The largest z-scores were obtained for 3T FLAIR. 3T and 9.4T quantitative T1 have similar effect sizes. Raw, non-z-scored data are shown in supplementary figure 4. (B) Horizontal profiles of different features, in bins with equal number of vertices in increasing distance from the centroid of the manually defined lesion label. The intensity features (FLAIR, T1, T2*, QSM) were sampled at 3 mm sub-pial depth. The topmost bar shows the fraction of vertices inside the manually defined lesion label in grayscale, the distance where the fraction is 0.5 is marked with a vertical dashed lined. The lesion extent appears similar for data acquired at both magnetic field strengths.

**Figure 5:**
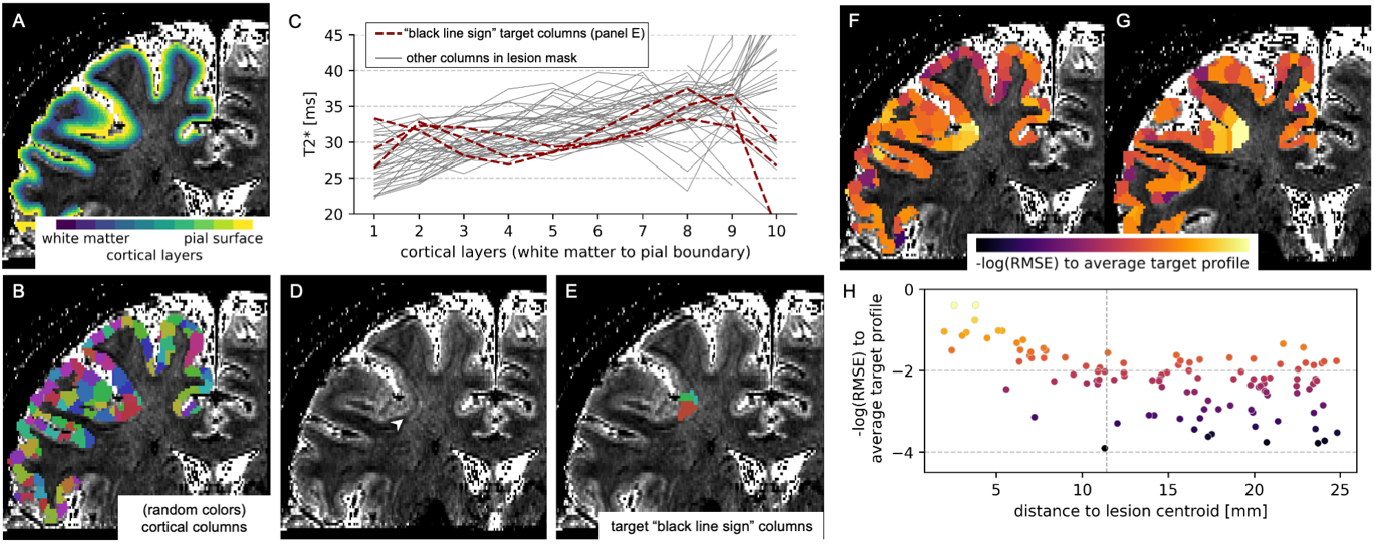
High resolution layer-wise analysis of T2* at 9.4T in a histologically verified FCD IIb. Cortical voxels were assigned to 10 equivolumetric cortical layers (A) and cortical columns with a volume of 40 mm3 each (B) using the laynii software suite. Profiles of T2* vs. cortical layer were extracted, (C) shows the profiles of all columns in the manual lesion mask. Columns overlapping with the visually apparent black line sign (white arrowhead in D, raw T2* map) were selected according to manually set criteria (see methods). These selected target columns’ profiles are colored in red in (C), whereas other columns’ profiles (within the lesion mask, but outside the black line sign area) are plotted in grey. In a next step, for each column, as a measure of similarity of its T2* profile to the average target profile, the log-transformed root mean square error (RMSE) was computed. These values are plotted for two coronal slices (F, G) and plotted against the distance from the lesion centroid (H). Columns within 5 mm distance from the lesion centroid are more similar to the target black line sign T2* profile, whereas columns in > 5 mm distance are less similar and exhibit higher variability.

We also inspected the data with a focus on horizontal lesion extent (i.e., parallel to the cortical surface), for both lesions the differences compared to controls were apparent for all features except for QSM, up to approximately 10-15 mm from the ROI centroid (see figure 4B). This aligns with the extent of the manually drawn lesion labels.

We further focused on the reduced T2* values mid-cortically in the centre of the FCD in patient P017 (see figure 5), which corresponds to the previously described “black line sign”^22,23^. We attempted to quantitatively describe this feature by defining a high-resolution cortical profile in the T2* maps with 0.375 mm in-plane resolution. A mid-cortical decrease in T2* was apparent in quantitative profiles of columns at the bottom of the sulcus (see figure 5 C-E). We observed a distance-dependent decrease of profile similarity to the average target profile from the center of the lesion to further outward (see figure 5 F-H). When applying the same target profile to the other FCD lesion (patient P026) without a visually apparent “black line sign”, we did not observe the same pattern (supplementary figure 5).

## Discussion

We present data from n=21 focal epilepsy patients and n=20 healthy controls scanned at 9.4T MRI and compare them to 3T MRI data in the same participants. Overall, epileptogenic lesions are visible at both field strengths. 9.4T MRI was able to recover features classically seen at 3T, some with increased detail due to higher resolution. Specifically, in one FCD, there was a visible and quantifiable intralesional microstructural heterogeneity (reduced intra-cortical T2* values, reflective of the “black line sign”) that we here aimed to quantify automatically. Lesions missed on 3T MRI and subsequently confirmed histologically were not present in this cohort; although such cases would be particularly informative for UHF-MRI analysis. This study provides new details on how UHF-MRI data can be used to quantitatively profile cortical pathology.

Research tools generating automatic predictions can increase the diagnostic yield for epileptogenic lesions, especially FCDs.^8^ These employ different methods, such as end-to-end deep learning^41,42^ or cortical surface-based morphometry.^11,12^ However, to date, none of these approaches has been optimized or evaluated for UHF-MRI data. In this study, we apply a surface-based reconstruction with preprocessing steps adapted for the MP2RAGE 9.4T data and compare quantitative features of FCDs across 9.4T and 3T MRI data. Depth-dependent changes and horizontal lesion spread that are distinctive for the lesion can be detected at both field strengths (figure 5). UHF-MRI enables optimized T2*-weighted imaging and quantitative mapping of T2* as well as susceptibility (QSM).^43^ In FCD IIb, a focal hypointensity on 7T T2*-weighted images in the center of the lesion has been termed “black line sign”.^22,23^ It has been found to be concordant with the seizure onset zone identified by invasive-EEG. Furthermore, resection of the respective region was associated with better post-surgical outcomes. Here, we observe a similar alteration on 9.4T T2* maps in one of two FCD IIb.

We also indicate that this pattern can be resolved quantitatively with high-resolution cortical profiling, which motivates future research to extract this feature automatically from UHF-MRI data. UHF-MRI may thus have a role in increasing the precision of the MRI-defined epileptogenic lesion and neurosurgical resection or ablation target.

In contrast to previous works that compared 7 T with 3T MRI, we could not identify new, previously undetected epileptogenic lesions using 9.4T MRI in this cohort. The diagnostic gain (i.e., fraction of MRI-negative cases with new relevant findings) of 7 T MRI has been reported as 84/205 (41%) in a systematic review of studies up to 2021^16^, and in more recent studies as: 3/8 (38%)^17^, 4/24 (17%)^18^, 4/21 (19%)^19^, 9/31 (29%)^15^, 9/60 (15%)^20^, 6/26 (23%)^21^. In a single reported case of 9.4T MRI in a 3T and 7 T negative focal epilepsy patient, no new lesion was identified.^26^ This discrepancy between previous studies and our data is likely due to methodological limitations. With 21 patients, although comparable to some of the cited studies, our cohort is relatively small. In addition, none of the 3T MRI negative patients recruited in this study later underwent a surgical resection and had clear histopathological findings, or an improved clinical outcome. This limits the statistical power to detect an improvement in diagnostic performance and the assessment of a possible gain in sensitivity.

In addition, at the time of data acquisition for this study, it was not feasible to acquire a standard FLAIR sequence at 9.4T due to SAR (specific absorption rate) limitations, i.e., due to safety aspects related to RF-induced tissue heating. However, FLAIR is one of the most relevant contrasts for detecting hippocampal sclerosis and FCDs.^10,44^ At 7 T MRI, FLAIR sequences can already be acquired, and FLAIR features, like the transmantle sign in FCD IIb, are important for identifying new lesions.^21^ This limits the current applicability of 9.4T compared to 7 T MRI. Finally, RF nonuniformity and susceptibility effects are more pronounced at 9.4T and can cause visible artefacts, especially in the lower temporal lobes (see supplementary figure 3). Parallel transmit methodology (pTx) has been introduced to mitigate these issues and improves diagnostic performance at 7 T.^15^ However, it was not yet possible to apply pTx pulses robustly at the time of data acquisition for this study.

A previous 3T MRI study measured R2* and QSM maps in 18 patients with histopathologically diagnosed FCD. They found decreased QSM χ values in FCD IIb most pronounced at a sub-pial sampling depth of 2 to 4 mm, but no changes in R2*.^45^ For the two FCD IIb lesions available here, we observed a different pattern: An increase in T2* and no changes in QSM values in the data obtained at 9.4T. This suggests altered T2* in FCD lesions rather than a difference in susceptibility. However, the interpretation is complicated by a recent observation that widespread alterations in qT2 can be observed in focal epilepsy patients also distant from the presumed seizure onset.^46^ An ex-vivo study using quantitative histology and 9.4T MRI described changes in T2 and T2* in an FCD IIa and changes in T1, T2 and T2* in an FCD IIb, which are attributed mainly to reduced myelination and altered neuronal density.^47^

In conclusion, imaging characteristics of FCDs at 9.4T parallel 3T findings. Quantitative feature profiles in T1 and T2*-weighted sequences differentiate lesional cortical and subcortical areas from homotopic areas in patients and homologue areas in healthy controls. High-resolution T2*-weighted imaging can provide additional diagnostic information, such as by identifying the “black line sign” that has previously been shown to be concordant with the seizure onset zone. This study does not demonstrate an increased diagnostic sensitivity using 9.4T MRI with respect to the detection of lesions in 3T MRI negative patients. Future work will include applying and adapting computational tools for automated lesion detection to the 9.4T MRI data.

## Data Availability

The healthy volunteer 9.4T MP2RAGE data has been published as part of a larger dataset [ref. 40]. It is currently not possible to make the patient imaging data openly accessible due to data protection regulations. The code used for analysis is available at https://github.com/ckronlage/3T9T_profiling_pub.

## Acknowledgements

We acknowledge the following funding: MINT-CS (Faculty of Medicine Tuebingen, Deutsche Forschungsgemeinschaft DFG) – CK. PRECISE.net (Else Kröner Fresenius Stiftung) – CK. de.NBI Cloud within the German Network for Bioinformatics Infrastructure (de.NBI) and ELIXIR-DE (Forschungszentrum Jülich and W-de.NBI-001, W-de.NBI-004, W-de.NBI-008, W-de.NBI-010, W-de.NBI-013, W-de.NBI-014, W-de.NBI-016, W-de.NBI-022) – CK. The Wellcome Trust – MR, KW. The Epilepsy Research Institute (P2208) – SA. EU-LACH Grant #16/T01-0118 – GEH. BMBF Grant 01GQ2101 – GEH, KS.

## Literature

1. Bien CG, Szinay M, Wagner J, Clusmann H, Becker AJ, Urbach H. Characteristics and Surgical Outcomes of Patients With Refractory Magnetic Resonance Imaging–Negative Epilepsies. Archives of Neurology. 2009; 66(12):1491–9.

2. Wang ZI, Alexopoulos AV, Jones SE, Jaisani Z, Najm IM, Prayson RA. The pathology of magnetic-resonance-imaging-negative epilepsy. Modern Pathology. 2013; 26(8):1051–8.

3. Téllez-Zenteno JF, Ronquillo LH, Moien-Afshari F, Wiebe S. Surgical outcomes in lesional and non-lesional epilepsy: A systematic review and meta-analysis. Epilepsy Research. 2010; 89(2):310–8.

4. Pichardo-Rojas D, Espinosa-Cantú CB, Valenzuela-Rangel AF, Choque-Ayala LC, Barrón-Lomelí A, Gutierrez-Herrera EA, et al. A comparative assessment of laser interstitial thermal therapy and open resective surgery for drug-resistant epilepsy: a meta-analysis of 3873 patients. Journal of Neurosurgery. 2025; 144(1):35–54.

5. Macdonald-Laurs E, Maixner WJ, Bailey CA, Barton SM, Mandelstam SA, Yuan-Mou Yang J, et al. One-Stage, Limited-Resection Epilepsy Surgery for Bottom-of-Sulcus Dysplasia. Neurology [Internet]. 2021 [cited 2026]; 97(2). Available from: https://www.neurology.org/doi/10.1212/WNL.0000000000012147

6. Zhao B, Zhang C, Wang X, Wang Y, Liu C, Mo J, et al. Sulcus-centered resection for focal cortical dysplasia type II: surgical techniques and outcomes. Journal of Neurosurgery. 2020; 135(1):266–72.

7. Najm I, Lal D, Alonso Vanegas M, Cendes F, Lopes-Cendes I, Palmini A, et al. The ILAE consensus classification of focal cortical dysplasia: An update proposed by an ad hoc task force of the ILAE diagnostic methods commission. Epilepsia. 2022; 63(8):1899–919.

8. Walger L, Schmitz MH, Bauer T, Kügler D, Schuch F, Arendt C, et al. A public benchmark for human performance in the detection of focal cortical dysplasia. Epilepsia Open. 2025; 10(3):778–86.

9. Adler S, Wagstyl K, Gunny R, Ronan L, Carmichael D, Cross JH, et al. Novel surface features for automated detection of focal cortical dysplasias in paediatric epilepsy. Neuroimage Clin. 2017; 14:18–27.

10. Hong S-J, Bernhardt BC, Caldairou B, Hall JA, Guiot MC, Schrader D, et al. Multimodal MRI profiling of focal cortical dysplasia type II. Neurology. 2017; 88(8):734–42.

11. Ripart M, Spitzer H, Williams LZJ, Walger L, Chen A, Napolitano A, et al. Detection of Epileptogenic Focal Cortical Dysplasia Using Graph Neural Networks: A MELD Study. JAMA Neurol [Internet]. 2025 [cited 2025];. Available from: https://jamanetwork.com/journals/jamaneurology/fullarticle/2830410

12. Spitzer H, Ripart M, Whitaker K, Napolitano A, De Palma L, De Benedictis A, et al. Interpretable surface-based detection of focal cortical dysplasias: a MELD study [Internet]. Neurology; 2021 [cited 2022]. Available from: http://medrxiv.org/lookup/doi/10.1101/2021.12.13.21267721

13. Pohmann R, Speck O, Scheffler K. Signal-to-noise ratio and MR tissue parameters in human brain imaging at 3, 7, and 9.4 tesla using current receive coil arrays. Magnetic Resonance in Medicine. 2016; 75(2):801–9.

14. Ladd ME, Bachert P, Meyerspeer M, Moser E, Nagel AM, Norris DG, et al. Pros and cons of ultra-high-field MRI/MRS for human application. Progress in Nuclear Magnetic Resonance Spectroscopy. 2018; 109:1–50.

15. Klodowski K, Zhang M, Jen JP, Scoffings DJ, Morris R, Lupson V, et al. Parallel transmit 7T MRI for adult epilepsy pre-surgical evaluation. Epilepsia. 2025; 66(7):2315–27.

16. van Lanen RHGJ, Colon AJ, Wiggins CJ, Hoeberigs MC, Hoogland G, Roebroeck A, et al. Ultra-high field magnetic resonance imaging in human epilepsy: A systematic review. NeuroImage: Clinical. 2021; 30:102602.

17. Bubrick EJ, Gholipour T, Hibert M, Cosgrove GR, Stufflebeam SM, Young GS. 7T versus 3T MRI in the presurgical evaluation of patients with drug-resistant epilepsy. Journal of Neuroimaging. 2022; 32(2):292–9.

18. Chen C, Xie J-J, Ding F, Jiang Y-S, Jin B, Wang S, et al. 7T MRI with post-processing for the presurgical evaluation of pharmacoresistant focal epilepsy. Ther Adv Neurol Disord. 2021; 14:17562864211021181.

19. Hangel G, Kasprian G, Chambers S, Haider L, Lazen P, Koren J, et al. Implementation of a 7T Epilepsy Task Force consensus imaging protocol for routine presurgical epilepsy work-up: effect on diagnostic yield and lesion delineation. J Neurol. 2024; 271(2):804–18.

20. van Lanen RHGJ, Uher D, Hoeberigs CMC, Hofman PAM, Santegoeds R, Wiggins C, et al. Value of ultra-high-field MRI in patients with drug-resistant focal epilepsy and negative 3T MRI (EpiUltraStudy): Diagnostic gain of 7T structural analysis. Epilepsia [Internet]. 2025 [cited 2025]; n/a(n/a). Available from: https://onlinelibrary.wiley.com/doi/abs/10.1111/epi.18682

21. Vecchiato K, Casella C, Dokumaci AS, Siddiqui A, Egloff A, Carney O, et al. Ultra-High Field 7T MRI in a Drug-Resistant Pediatric Epilepsy Cohort: Image Comparison and Radiologic Outcomes. Neurology. 2025; 105(5):e213921.

22. Bartolini E, Cosottini M, Costagli M, Barba C, Tassi L, Spreafico R, et al. Ultra-High-Field Targeted Imaging of Focal Cortical Dysplasia: The Intracortical Black Line Sign in Type IIb. AJNR Am J Neuroradiol. 2019; :ajnr;ajnr.A6298v1.

23. Tang Y, Blümcke I, Su T-Y, Choi JY, Krishnan B, Murakami H, et al. Black Line Sign in Focal Cortical Dysplasia IIB: A 7T MRI and Electroclinicopathologic Study. Neurology. 2022; 99(6):e616–26.

24. Boulant N, Mauconduit F, Gras V, Amadon A, Le Ster C, Luong M, et al. In vivo imaging of the human brain with the Iseult 11.7-T MRI scanner. Nat Methods. 2024; 21(11):2013–6.

25. Martin P, Hagberg G, Loureiro J, Bause J, Lerche H, Focke N, et al. Can 9.4 T MRI improve lesion visualization in epilepsy patients? In: ESMRMB 2016, 33rd Annual Scientific Meeting, Vienna, AT, September 29 – October 1: Abstracts, Saturday [Internet]. 2016 [cited 2025]. p. S347–8. Available from: http://link.springer.com/content/pdf/10.10072Fs10334-016-0570-3.pdf

26. Van Lanen RHGJ, Uher D, Tse DHY, Steijvers E, Colon AJ, Jansen JFA, et al. In vivo 9.4 Tesla MRI of a patient with drug-resistant epilepsy: Technical report. Acta Neurochir. 2025; 167(1):18.

27. van Lanen RHGJ, Wiggins CJ, Colon AJ, Backes WH, Jansen JFA, Uher D, et al. Value of ultra-high field MRI in patients with suspected focal epilepsy and negative 3 T MRI (EpiUltraStudy): protocol for a prospective, longitudinal therapeutic study. Neuroradiology. 2022; 64(4):753–64.

28. Shajan G, Kozlov M, Hoffmann J, Turner R, Scheffler K, Pohmann R. A 16-channel dual-row transmit array in combination with a 31-element receive array for human brain imaging at 9.4 T. Magnetic Resonance in Medicine. 2014; 71(2):870–9.

29. Marques JP, Kober T, Krueger G, van der Zwaag W, Van de Moortele P-F, Gruetter R. MP2RAGE, a self bias-field corrected sequence for improved segmentation and T1-mapping at high field. NeuroImage. 2010; 49(2):1271–81.

30. Hagberg G, Bause J, Ethofer T, Ehses P, Dresler T, Herbert C, et al. Whole brain MP2RAGE-based mapping of the longitudinal relaxation time at 9.4T. NeuroImage. 2017; 144:203–16.

31. Hagberg GE, Eckstein K, Tuzzi E, Zhou J, Robinson S, Scheffler K. Phase-based masking for quantitative susceptibility mapping of the human brain at 9.4T. Magnetic Resonance in Medicine. 2022; 88(5):2267–76.

32. Loureiro JR, Himmelbach M, Ethofer T, Pohmann R, Martin P, Bause J, et al. In-vivo quantitative structural imaging of the human midbrain and the superior colliculus at 9.4T. NeuroImage. 2018; 177:117–28.

33. O’Brien KR, Kober T, Hagmann P, Maeder P, Marques J, Lazeyras F, et al. Robust T1-Weighted Structural Brain Imaging and Morphometry at 7T Using MP2RAGE. PLOS ONE. 2014; 9(6):e99676.

34. Tustison NJ, Avants BB, Cook PA, Zheng Y, Egan A, Yushkevich PA, et al. N4ITK: Improved N3 Bias Correction. IEEE Transactions on Medical Imaging. 2010; 29(6):1310–20.

35. Fischl B. FreeSurfer. NeuroImage. 2012; 62(2):774–81.

36. Hoopes A, Mora JS, Dalca AV, Fischl B, Hoffmann M. SynthStrip: skull-stripping for any brain image. NeuroImage. 2022; 260:119474.

37. Focke NK, Symms MR, Burdett JL, Duncan JS. Voxel-based analysis of whole brain FLAIR at 3T detects focal cortical dysplasia. Epilepsia. 2008; 49(5):786–93.

38. Greve DN, Van der Haegen L, Cai Q, Stufflebeam S, Sabuncu MR, Fischl B, et al. A Surface-based Analysis of Language Lateralization and Cortical Asymmetry. Journal of Cognitive Neuroscience. 2013; 25(9):1477–92.

39. Huber L (Renzo), Poser BA, Bandettini PA, Arora K, Wagstyl K, Cho S, et al. LayNii: A software suite for layer-fMRI. NeuroImage. 2021; 237:118091.

40. Mahler L, Steiglechner J, Bender B, Lindig T, Ramadan D, Bause J, et al. UltraCortex: Submillimeter Ultra-High Field 9.4 T Brain MR Image Collection and Manual Cortical Segmentations [Internet]. arXiv; 2025 [cited 2025]. Available from: http://arxiv.org/abs/2406.18571

41. Gill RS, Lee H-M, Caldairou B, Hong S-J, Barba C, Deleo F, et al. Multicenter Validation of a Deep Learning Detection Algorithm for Focal Cortical Dysplasia. Neurology. 2021; : 10.1212/WNL.0000000000012698.

42. Kersting LN, Walger L, Bauer T, Gnatkovsky V, Schuch F, David B, et al. Detection of focal cortical dysplasia: Development and multicentric evaluation of artificial intelligence models. Epilepsia [Internet]. [cited 2025]; n/a(n/a). Available from: https://onlinelibrary.wiley.com/doi/abs/10.1111/epi.18240

43. Budde J, Shajan G, Hoffmann J, Uğurbil K, Pohmann R. Human imaging at 9.4 T using T2*-, phase-, and susceptibility-weighted contrast. Magnetic Resonance in Medicine. 2011; 65(2):544–50.

44. Pastore LV, Sudhakar SV, Mankad K, De Vita E, Biswas A, Tisdall MM, et al. Integrating standard epilepsy protocol, ASL-perfusion, MP2RAGE/EDGE and the MELD-FCD classifier in the detection of subtle epileptogenic lesions: a 3 Tesla MRI pilot study. Neuroradiology [Internet]. 2024 [cited 2025];. Available from: 10.1007/s00234-024-03488-8

45. Lorio S, Sedlacik J, So P-W, Parkes HG, Gunny R, Löbel U, et al. Quantitative MRI susceptibility mapping reveals cortical signatures of changes in iron, calcium and zinc in malformations of cortical development in children with drug-resistant epilepsy. NeuroImage. 2021; 238:118102.

46. Casella C, Vecchiato K, Cromb D, Guo Y, Winkler AM, Hughes E, et al. Widespread, depth-dependent cortical microstructure alterations in pediatric focal epilepsy. Epilepsia. 2024; 65(3):739–52.

47. Reeves C, Tachrount M, Thomas D, Michalak Z, Liu J, Ellis M, et al. Combined Ex Vivo 9.4T MRI and Quantitative Histopathological Study in Normal and Pathological Neocortical Resections in Focal Epilepsy. Brain Pathology. 2016; 26(3):319–33.

