## Supplementary Material for "9.4 Tesla MRI in focal epilepsy patients with high-resolution surface-based profiling of focal cortical dysplasias"

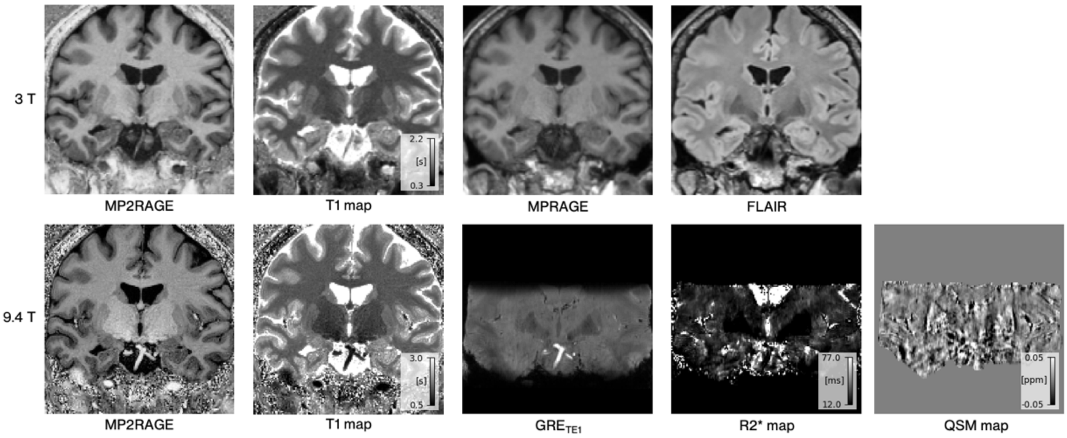


Supplementary figure 1: Coronal slices patient P009 with suspected right hippocampal sclerosis. Comparison of 3T data (top row, coregistered and resampled to the 9.4T volume with cubic interpolation) and 9.4T data (bottom row). Volume loss of the right hippocampus and corresponding enlargement of the temporal horn of the lateral ventricle can be appreciated in the 3T images. There is no clear T2/FLAIR hyperintensity. Right hippocampal volume loss is also clearly visible in the 9.4T image, with more sharply defined borders. 9.4T GRE and derived maps (T2*, QSM) do appear to show a contrast in this area and are affected by artefacts in proximity to the skull base.

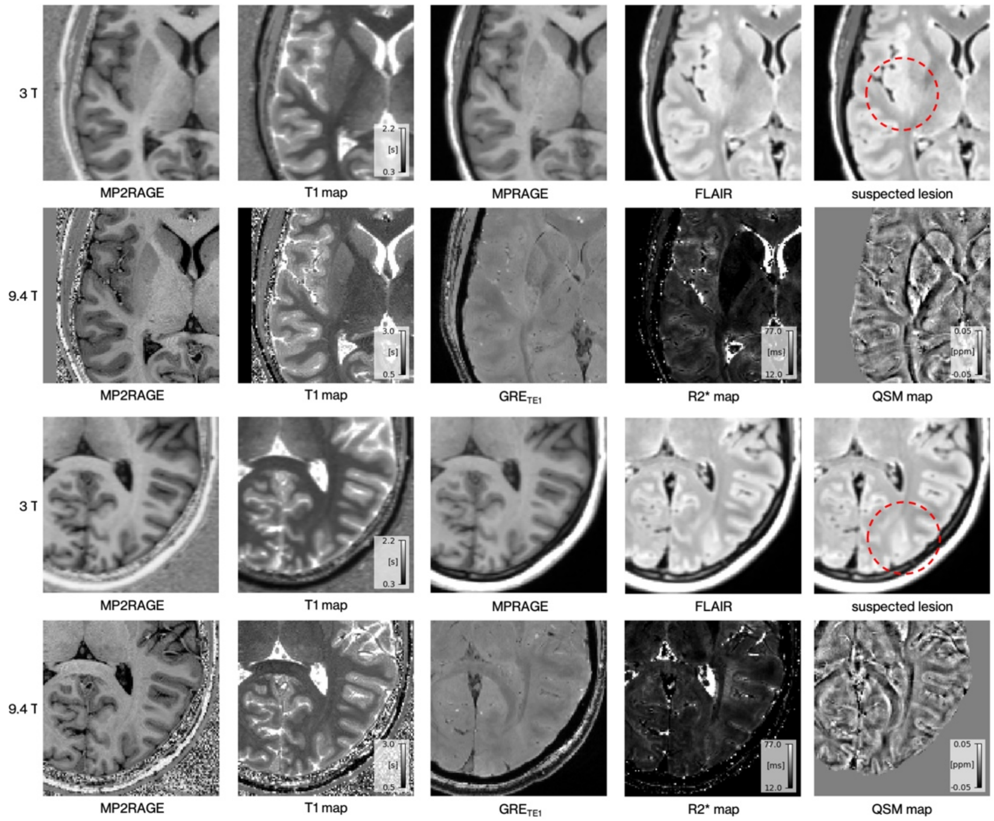


Supplementary figure 2: Axial slices of the two suspected FCD in patient P016 where no seizures could be recorded in video-EEG and a differential diagnosis of non-epileptic seizures was discussed (also rendering the diagnosis of FCDs uncertain). Top rows: An asymmetrical T2/FLAIR hyperintensity is visible in the posterior right insular cortex, in the 9.4T GRE a similar finding can be suspected, the T1-weighted sequences reveal no clear abnormality. Bottom rows: A blurring of the GM/WM boundary and subcortical FLAIR-hyperintensity is visible in the occipital sulcus in the 3T data.

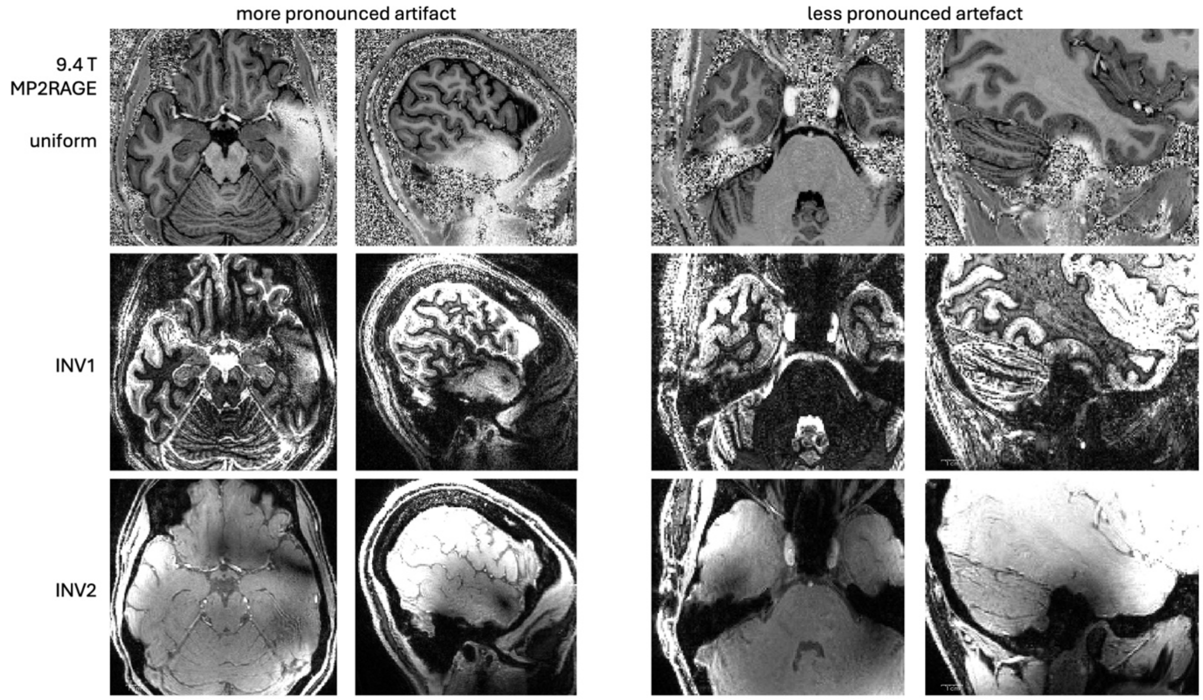


Supplementary figure 3: Artefacts in the temporal lobes in the 9.4T MP2RAGE data. Left: Example of a more pronounced artefact (as present in 3/41 subjects in total). Right: Example of a less pronounced artefact as present in all of the subjects. Top row: MP2RAGE uniform volumes, middle row: MP2RAGE INV1, bottom row: MP2RAGE INV2. These findings are likely mainly caused by to transmit field inhomogeneities.

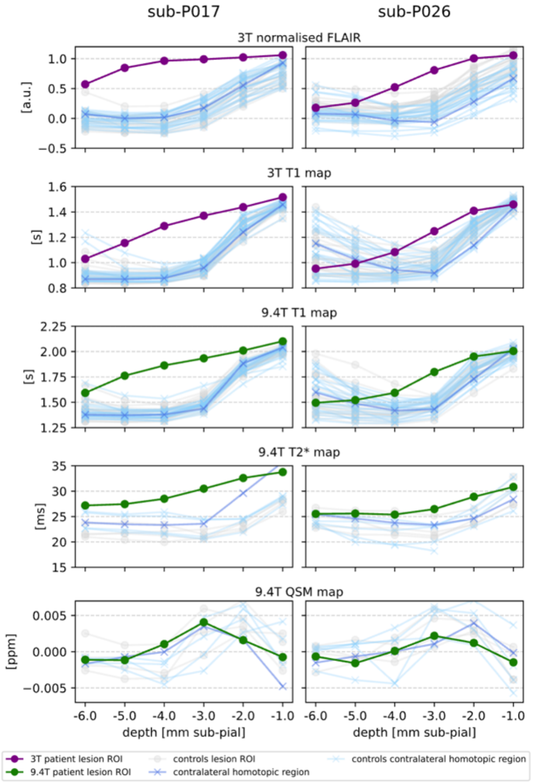


Supplementary figure 4: Vertical profiles of two histologically verified FCDs across magnetic field strengths of 3T and 9.4T. For each feature and depth, the raw feature value in the lesion ROI is plotted. Also see figure 4.

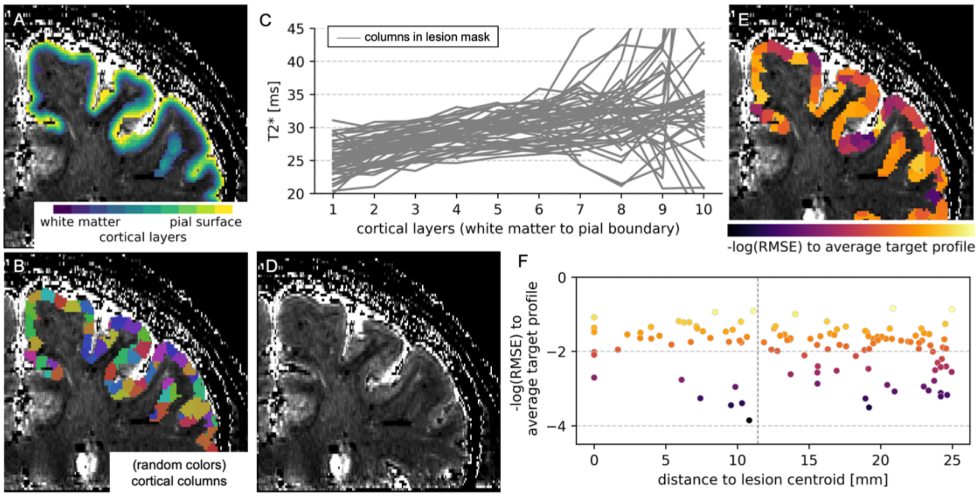


Supplementary figure 5: High resolution layer-wise analysis of T2* at 9.4T in a histologically verified FCD IIb in patient P026. Cortical voxels were assigned to 10 equivolumetric cortical layers (A) and cortical columns with a volume of 40 mm3 each (B) using the laynii software suite. Profiles of T2* vs. cortical layer were extracted, (C) shows the profiles of all columns in the manual lesion mask. In this case, there were no columns matching the manually set criteria (see methods) for the “black line sign” feature. (D) shows the T2* map without overlays. We assessed the similarity of P026 column T2* profiles to the target profile derived from patient P017. The log-transformed root mean square error (RMSE) is plotted overlaid onto the respective columns (E) and against the distance from the lesion centroid (F). In this case, although individual columns have a higher similarity score, there is no consistent distance-dependent pattern as in P017.
